# Measles-rubella surveillance and its facilitators and barriers in Bagmati province, Nepal: A mixed methods study

**DOI:** 10.1101/2025.10.09.25337666

**Authors:** Bipsana Shrestha, Surakshya Kunwar, Alina Upreti, Kshitij Karki, Paras Pangeni, Poonam Subedi, Leela Khanal, Rajeev Shrestha, Dipesh Shrestha, Bhim Singh Tinkari, Shyam Raj Upreti

## Abstract

**Objective:** This study aimed to assess the status of measles-rubella surveillance and understand the facilitators and barriers for the measles-rubella surveillance activities in selected municipalities of selected districts of Bagmati province of Nepal.

**Design:** This is a convergent parallel mixed method study, incorporating both quantitative and qualitative approaches, was conducted in eight municipalities of the three districts (Sindhupalchok, Makwanpur, and Chitwan) of Bagmati province, Nepal.

**Setting:** Three districts from Bagmati Province were purposively selected to ensure ecological representation: Sindhupalchok (mountain), Makwanpur (hill), and Chitwan (Terai). Eight municipalities, including both rural and urban areas, were selected from the three districts. The health facilities included in the study were: a) All the health facilities that are part of the WHO reporting unit (RU) for Vaccine-Preventable Disease (VPD) surveillance b) purposively selected health facilities outside the WHO RU. Health facilities within each municipality were selected, ensuring representation of government hospitals, private hospitals, Primary Health Care Centers (PHCC), and Health Posts (HP).

**Participants:** From each selected district, one District Health Office Chief and one immunization and surveillance focal person were purposively recruited for Key Informant Interviews (KIIs), along with one health section chief from each selected municipality. A total of 14 KIIs were conducted with district and municipal stakeholders. For in-depth interviews (IDIs), two WHO-SMOs overseeing VPD surveillance and immunization activities in these districts, along with a VPD surveillance focal person from each selected reporting and non-reporting health facility, were purposively selected.

**Results:** The key strategies to achieve measles elimination, that is the MR2 coverage of 95%, was not achieved in most of the municipalities. The main barriers to effective VPD surveillance among others included limited awareness of VPDs under surveillance, poor understanding of the measles elimination target, and lack of clarity on standard case definitions for suspected measles-rubella cases. The major facilitators for effective VPD surveillance included awareness on the importance of VPD surveillance in measles elimination, availability of SCD posted in all reporting health facilities, and timely case notification and investigation by RUs.

**Conclusions:** Although, national immunization programme (NIP) is a priority 1 (P1) programme of the Government of Nepal, one of the key strategies (95% MR2 coverage) to achieve measles elimination was not achieved in most of the municipalities. This study suggests that measles-rubella surveillance in the study sites can be strengthened through capacity building of health workers in VPD surveillance, ensuring the availability of the VPD surveillance field guide, availability of the dedicated focal person, emphasizing in data analysis and utilization of surveillance data.

**Strengths and limitations of this study:** One of the major strengths of this study is the selection of districts from all three ecological regions of the country as it helps to ensure geographical coverage. It helps to increase the relevance of the results across similar ecological regions in Nepal. Similarly, our study also included both WHO reporting and non-reporting health facilities ranging different types of health facilities such as government hospitals, private hospitals, primary health care centers and health posts which helps to cover broader perspective on surveillance system. Likewise, wide-range of stakeholders (district health officers, municipal health staff, WHO-SMOs, and health facility focal persons) involvement in this study provides multiple viewpoints on facilitators and barriers of surveillance. Furthermore, this study included both rural as well as urban municipalities exploring the perspectives of focal persons from those settings.

Despite all these strengths, there are also some limitations of this study, and one of them is purposive sampling of districts, municipalities and participants which may have introduced selection bias and which may also limit the generalizability to all parts of Nepal. Additionally, the study was restricted to three districts of only one province of Nepal (Bagmati), therefore, it may not capture nation-wide variance in measles-rubella surveillance. Reporting biases is also one of the potential limitations of this study especially in case of qualitative data collection from district health officers, municipal health staff, WHO-SMOs, and health facility focal persons because they may have provided socially desirable responses affecting the actual findings of KIIs and IDIs.

## Introduction

Measles is a highly contagious VPD that can cause serious disease and death. It remains a major cause of morbidity and mortality worldwide, with an estimated over 9 million cases and 136,000 deaths every year.^1^ Worldwide, millions of children did not receive their vaccinations during the COVID-19 pandemic, which led to an 18% rise in estimated measles cases and a 43% rise in estimated measles deaths in 2022 compared to 2021.^2^ However, in Nepal, the number of suspected measles cases declined from 1417 in 2021 to 1108 in 2022 following the Measles Rubella Supplementary Activities (MR SIA) in 2020 alongside routine immunization.^3^ In 2023, Nepal recorded a total of 962 confirmed measles cases and 14 confirmed measles outbreaks.^3^ Global measles and rubella strategic plan 2012-2020 includes high vaccination coverage with two doses of measles- and rubella-containing vaccines as one of its key strategies to eliminate measles.^4^ However, millions of children are unvaccinated or under-vaccinated, which has led to large measles outbreaks in every region of the world.^5^ In 2023, 83% of children had received the one dose of measles-containing vaccine, and 74% of children received two doses of measles vaccine worldwide.^6^ Several factors that contribute to being unvaccinated or under-vaccinated are driven by a complex mix of structural, social, and behavioral factors, which change with context and over time.^7^

In 2013, member states of the World Health Organization (WHO) South-East Asia Region (SEAR) adopted the goal of measles elimination and rubella and congenital rubella syndrome control by 2020.^8^ In 2019, SEAR member states declared a revised goal of eliminating both measles and rubella by 2023.^9^ Several challenges, including immunity gaps, suboptimal sensitivity of surveillance, inadequate outbreak response and preparedness, funding gaps, and the negative effects of the COVID-19 pandemic on immunization programs threaten achievement of the 2023 target.^10-11^ In 2023, only eight countries achieved the surveillance indicator for the target discarded rate of ≥2 per 100,000 population.^12^ A regional consultation meeting was held on resetting the target date for achieving the goal of measles and rubella elimination in the WHO South-East Asia Region in March 2023. The meeting has set a revised target year for measles elimination at 2026.^13^

Nepal along with other member states of the WHO SEAR in 2019, committed to eliminate measles-rubella by 2023 by implementing the ‘Strategic plan for measles and rubella elimination in WHO South-East Region:2020-2024’.^14^ Following the impact of the pandemic on the national immunization program and services, Nepal has now revised its target to achieve elimination by 2026 and has prepared the National Measles Rubella Elimination Roadmap.^15^ In Nepal, the national immunization schedule has included one dose of the measles vaccine at 9 months since 1990 and a second dose of MR at 15 months since 2015. The national coverage of MR1 was 97% and MR2 was 95% according to the Annual Health Report 2022/2023.^16^

High vaccine coverage and effective surveillance systems are indispensable to achieve measles elimination. In addition to these, timely and high-quality laboratory testing plays a supportive role in efforts to control and eliminate measles and rubella.^17^ In Nepal, all the collected samples from suspected cases are transported to WHO-accredited (National Public Health Laboratory, (NPHL) Kathmandu) and BP Koirala Institute of Health Sciences (BPKIHS), Dharan for laboratory confirmation by ELISA test. Shipping samples from hard-to-reach areas within 72 hours under a reverse cold chain is particularly challenging in a geographically diverse country like Nepal.^18^ The delay in sample transportation may result in significant delays in outbreak confirmation and the initiation of appropriate outbreak response measures. Although Nepal has achieved all the targeted measles-rubella surveillance performance indicators at the national level by the year 2023, the percentage of IgM results reported to public health authorities by the laboratory within 4 days of specimen receipt is 71% against the target 80%.^19^ The large number of clinically compatible cases reported in the case-based surveillance system highlights a failure to collect specimens for laboratory confirmation due to issues with timely specimen collection and transport to the National Public Health Laboratory at Kathmandu.^20^

Given these challenges, the use of simplified diagnostic tools could greatly improve the timeliness and effectiveness of measles testing, enabling prompt outbreak-response supplementary immunization activities (SIAs).^21^ This pilot study aims to identify the optimal deployment and testing strategy for Measles RDTs. Prior to piloting the deployment of Measles RDTs, a baseline assessment was conducted to evaluate the current measles disease status and surveillance performance. The results of this assessment are critical for: (1) determining the status of measles-rubella surveillance prior to Measles RDT deployment and (2) facilitating ongoing monitoring of improvements in surveillance and case detection rates.

## Materials and methods

### Study design and setting

A convergent parallel mixed method study, incorporating both quantitative and qualitative approaches, was conducted in eight municipalities of the three districts (Sindhupalchok, Makwanpur, and Chitwan) of Bagmati province, Nepal. A quantitative study was conducted to assess the measles-rubella immunization coverage, measles-rubella surveillance performance in selected municipalities through secondary data collection and availability of the necessary logistics and status of weekly reporting for suspected measles-rubella cases at the RU health facilities through observation checklist. In parallel, a qualitative approach was utilized to understand the facilitators and barriers of the measles-rubella surveillance system through key informant interviews with district and municipal stakeholders, and IDIs with WHO-Surveillance Medical Officers (WHO-SMOs), and immunization and surveillance focal personnel from health facilities in three districts of Bagmati Province, Nepal.

Three districts from Bagmati Province were purposively selected to ensure ecological representation: Sindhupalchok (mountain), Makwanpur (hill), and Chitwan (Terai). Eight municipalities, including both rural and urban areas, were selected from the three districts. The health facilities included in the study were: a) All the health facilities that are part of the WHO RU for VPD surveillance b) purposively selected health facilities outside the WHO RU. Health facilities within each municipality were selected, ensuring representation of government hospitals, private hospitals, Primary Health Care Centers (PHCC), and Health Posts (HP).

| District | Municipalities | Municipality type |
| --- | --- | --- |
| Sindhupalchok (Mountain) | Helambu rural municipality | Rural |
|  | Barhabise municipality | Urban |
| Makwanpur (Hill) | Hetauda sub-metropolitan | Urban |
|  | Bhimphedi rural municipality | Rural |
|  | Raksirang rural municipality | Rural |
| Chitwan (Terai) | Bharatpur metropolitan city | Urban |
|  | Ichchhakamana rural municipality | Rural |
|  | Kalika municipality | Rural |

### Sampling and sample size

From each selected district, one District Health Office Chief and one immunization and surveillance focal person were purposively recruited for Key Informant Interviews (KIIs), along with one health section chief from each selected municipality. A total of 14 KIIs were conducted with district and municipal stakeholders. For IDIs, two WHO-SMOs overseeing VPD surveillance and immunization activities in these districts, along with a VPD surveillance focal person from each selected RUs and non-reporting units (NRUs), were purposively selected. A total of 37 IDIs were conducted, with the sample size determined based on the principle of saturation.

**Figure 1.**
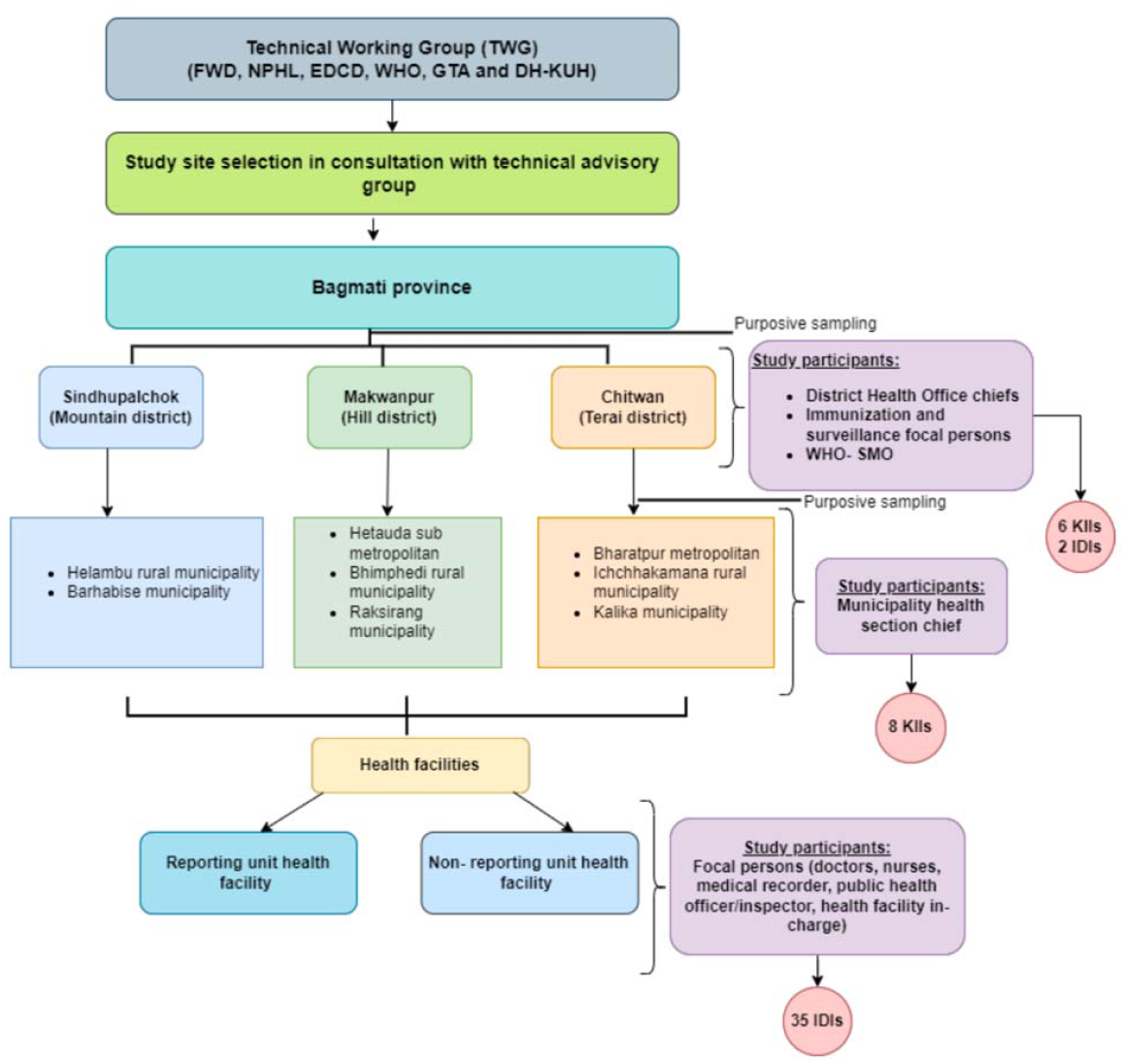
Sampling and sample size.

### Data collection tool

We used a structured matrix to collect quantitative data on three-year trend (2021-2023) of immunization (DPT1, DPT3, MR1, MR2) coverage and measles-rubella surveillance performance containing indicators for case detection, case notification and investigations, timeliness and completeness of VPD weekly reporting and measles-rubella outbreak. Further, an observation checklist was also used to assess the availability of the necessary logistics and status of weekly reporting for suspected measles-rubella cases at the RU health facilities.

For qualitative data collection, a semi-structured interview guide was used which was developed based on the literature review by consulting with experts of technical working group (TWG) of the project. The interview guides were based on the following themes like understanding on VPD surveillance, measles-rubella surveillance core functions, outbreak and response, measles-rubella surveillance supportive functions, experience and willingness of Rapid Diagnostic Test utilization.

### Data collection technique

For the quantitative data, a secondary data collection method was used to collect the data on immunization coverage and measles-rubella surveillance performance. Immunization coverage data were taken from health management information system (HMIS) of Government of Nepal. Secondary data for measles-rubella surveillance performance indicators were collected from the WHO-SMO of WHO-IPD field office of selected sites. A primary data collection method was used for observation to assess the availability of the necessary logistics and status of weekly reporting for suspected measles-rubella cases at the RU health facilities.

For qualitative data collection, the team conducted interviews with stakeholders from district and municipality, surveillance focal persons at respective health facilities and WHO-SMOs at their office. All interviews were carefully scheduled to avoid interfering with their work hours and patient care and were audio-recorded with participants’ consent. Each interview lasted between 40 to 50 minutes. The qualitative data collection was completed over one month, in June 2024. Before initiating the data collection, the tools were pre-tested following an iterative process in Lalitpur district of Lalitpur municipality (one of districts of Bagmati province). After the pretesting of tools, certain revisions were made to the data collection tools. The pre-tested data were not included in the data analysis of the study. Data were collected by trained research assistants with a qualification of bachelor’s degree in public health. Before deployment in the field, the assistants received four-day training on data collection techniques and ethical considerations.

### Data analysis

For the quantitative data, the data on immunization coverage and measles-rubella surveillance performance by three-year trend (2020-2023) were presented in frequency and percentage using tables and graphs as per the municipalities. The observation checklist which includes availability of the necessary logistics and status of weekly reporting for suspected measles-rubella cases in RU health facilities were presented as frequency and percentage.

For the qualitative interviews, to ensure anonymity, we assigned an alpha-numeric code to each participant and de-identified their transcripts. The IDIs and KIIs were audio recorded, transcribed into Nepali and translated into English language. The quality of the data was checked by reading the Nepali transcripts while listening to the audio files. An analytical framework of deductive codes was developed to reduce and summarize the data. A transcript was independently coded and compared among two researchers and their coding schemes were discussed to increase the rigor of analysis. A thematic analysis was done. Initially a deductive codebook was developed based on the interview guides, anticipated responses from the literature review, and pre-tested data (21). The analysis was done using Dedoose software version 9.2.0.12. The intercoder reliability (ICR) was checked and a score was calculated which was 9.59.

### Patient and public involvement

Participants were not involved in the design, conduct, reporting and dissemination plans of our research.

## Results

### Quantitative findings

Out of eight municipalities selected, the highest number of population of under 5 years of age was found to be in Bharatpur Metropolitan city (23973) and the lowest in Helambu rural municipality. The number of RUs was also found to be in Bharatpur Metropolitan city (8). Helambu, Barabise and Raksirang municipalities has single RUs in each of them.

**Table 1.** Selected municipalities and its demographic characteristics (n=8)

| Municipality Name | Total Population |  | Total population by age (under-5 years) | Total population by 15 months | No. of health facilities by type |  |  |  |  | No.of RUs |
| --- | --- | --- | --- | --- | --- | --- | --- | --- | --- | --- |
|  | Male | Female |  |  | Govt.h ospital | PHCC | HP/ CHU | UHC/ BHC | Private |  |
| Helambu Rural Municipality | 8783 | 8714 | 1286 | 324 | 0 | 1 | 7 | 2 | 0 | 1 |
| Barhabise Municipality | 12030 | 12079 | 1601 | 396 | 0 | 1 | 8 | 5 | 1 | 1 |
| Hetauda Sub-metropolitan City | 95626 | 97950 | 12860 | 2631 | 5 | 0 | 7 | 15 | 5 | 4 |
| Bhimphedi Rural Municipality | 10693 | 10823 | 1781 | 449 | 0 | 1 | 7 | 4 | 1 | 2 |
| Raksirang Rural Municipality | 13206 | 12790 | 3143 | 773 | 0 | 0 | 5 | 0 | 0 | 1 |
| Bharatpur Metropolitan City | 178726 | 190542 | 23973 | 5800 | 28 | 1 | 14 | 16 | 20 | 8 |
| Kalika Municipality | 25508 | 26656 | 4607 | 2924 | 0 | 1 | 3 | 7 | 0 | 2 |
| Ichchhakamana Rural Municipality | 14015 | 13628 | 1313 | 857 | 0 | 0 | 10 | 3 | 0 | 1 |
| **Total** |  |  |  |  | **33** | **5** | **61** | **52** | **27** | **20** |

The vaccine coverage of MR1 was highest in Barabise in 2020-2021 and MR2 was highest in Hetauda. Likewise, in 2021-2022, the vaccine coverage of both MR1 and MR2 was highest in Barabise. In 2022-2023, coverage of these vaccines was highest in Hetauda.

**Table 2.**
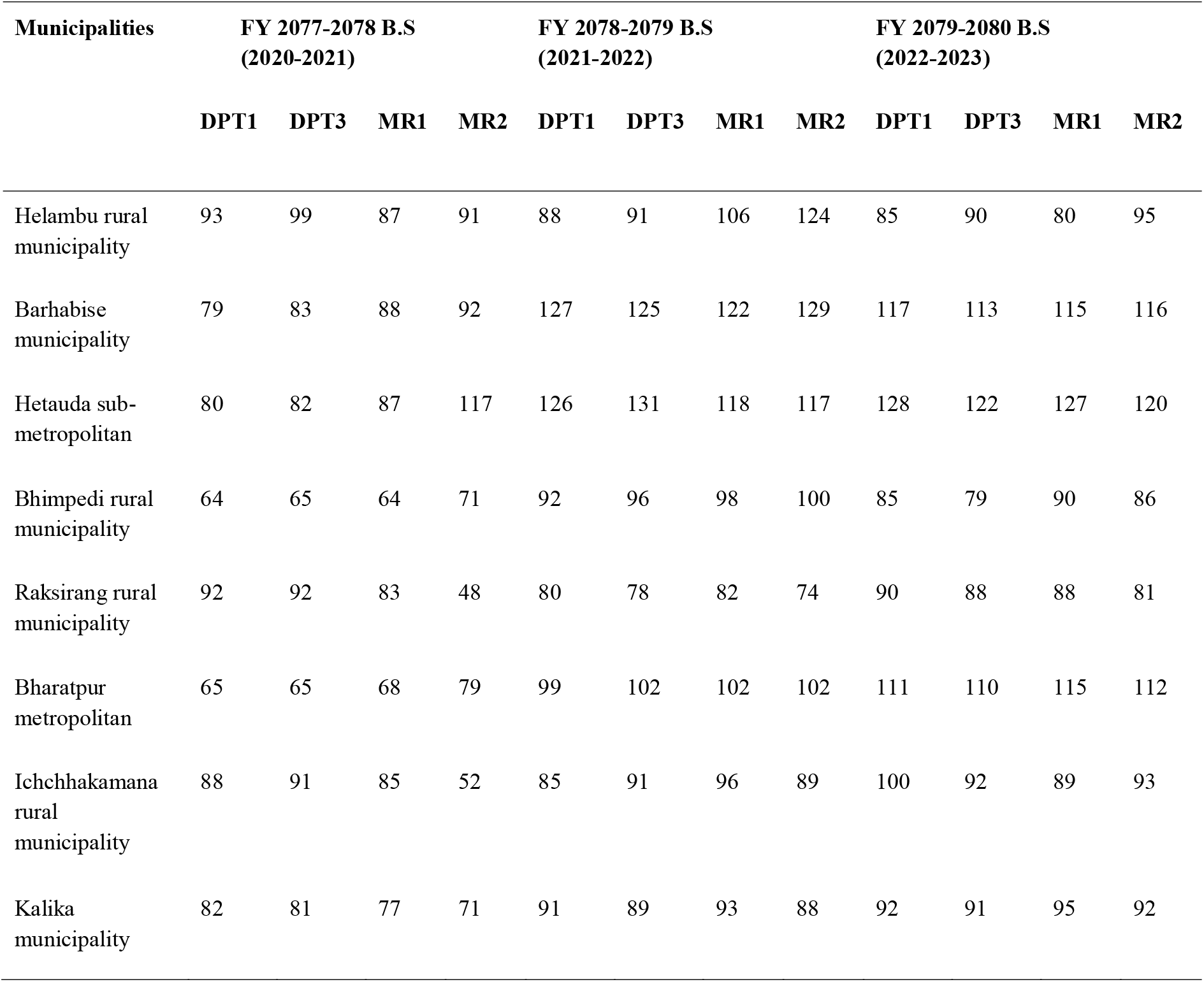
Vaccination coverage in the study sites by three years.

Table 3 shows the findings from the observation checklist that was conducted at the RU health facilities to assess the availability of the guidelines, standard operating procedures, logistics and supplies, completeness and timeliness of weekly reporting for suspected measles-rubella cases. VPD surveillance guidelines were not available in any health facilities. None of them had standard operating procedure for case investigation. During the observation, 90% of the health facilities (18 out of 20) had posted the Standard Case Definition (SCD) for measles and rubella cases. For the Case Investigation Forms (CIF), 80% of health facilities had these forms. In terms of VPD register availability, 95% of health facilities had these registers. 85% were equipped with sufficient specimen collection materials, including syringes, plain test tubes, and gloves as well as laboratory laboratory supplies such as lab unit, lab personnel, centrifuge equipment, cryo vial, refrigerator, cold box. In the last six months, 95% of health facilities submitted complete weekly reports whereas only 11 out of 20 health facilities submitted their weekly reports on time. Regarding the analysis of the surveillance data, none of the health facilities have done any analysis of the surveillance data.

**Table 3.**
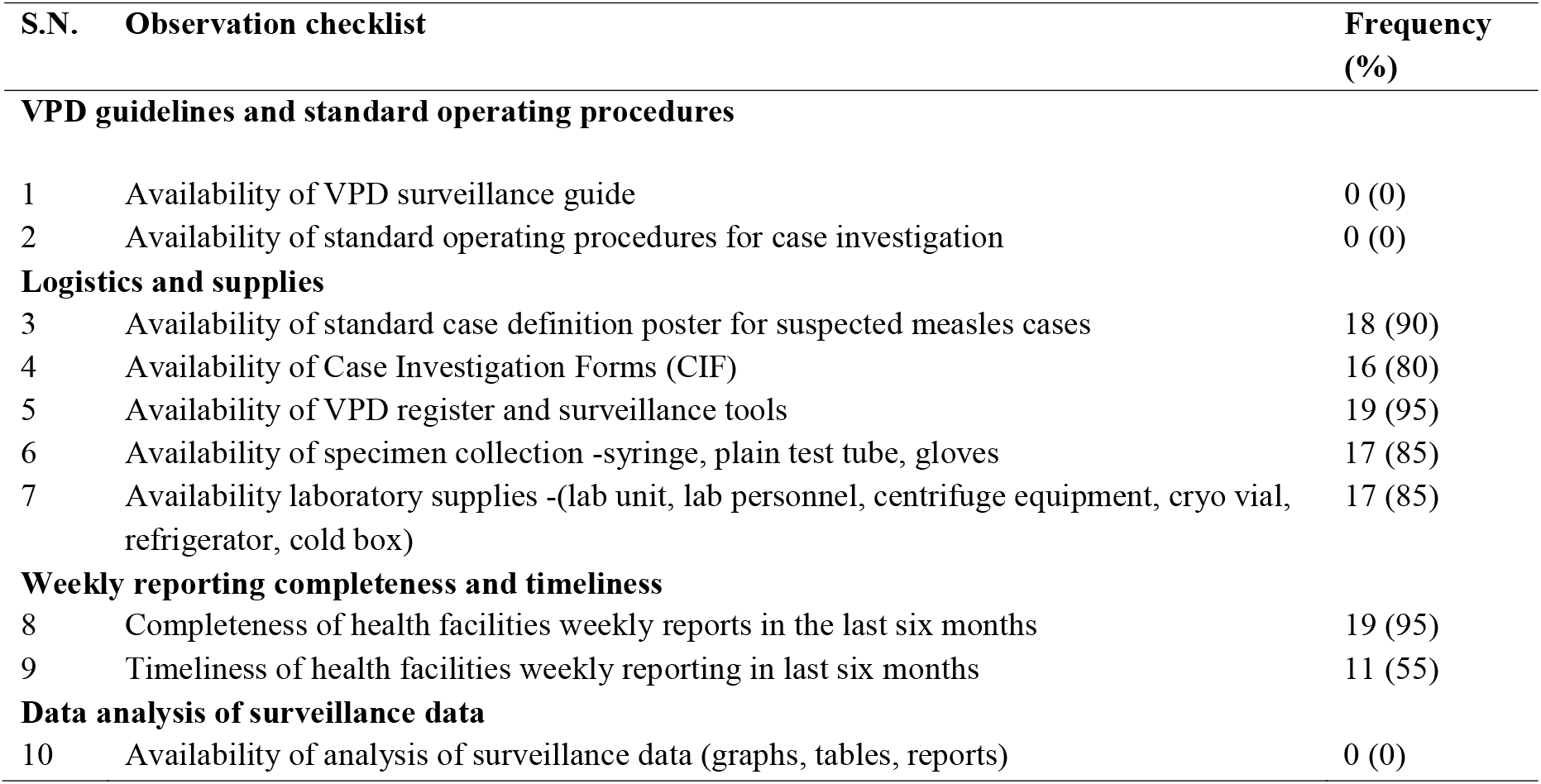
Observation checklist to assess the availability of the necessary logistics and status of weekly reporting.

### Qualitative findings

#### Theme 1: Understanding on VPD surveillance

Most of the respondents from all levels mentioned measles as one of the VPDs under current VPD surveillance. However, only a few could correctly name all four VPDs included in the surveillance: measles-rubella, neonatal tetanus, Japanese encephalitis, and acute flaccid paralysis. Some respondents mentioned diseases such as yellow fever, typhoid, tuberculosis, hepatitis B, and human papillomavirus (HPV), which are not part of the current VPD surveillance.

Only one fourth of the respondents were aware about the measles elimination target year set by the government. However, some including a few from NRUs were uncertain about it.

> *“I don’t know much about the measles elimination target year set by the government*.*”*
>
> ***-Municipal level respondent, Sindhupalchok KII***

Despite differing levels of understanding on the measles elimination target year, nearly all respondents except a few from NRUs mentioned that VPD surveillance plays a crucial role in measles elimination. They reported that surveillance is critical for early case detection, identifying the source of infection, forecasting potential outbreak areas, and planning for outbreak response.

> *“I do not have much idea on the importance of surveillance in measles elimination*.*”*
>
> ***-NRU respondent, Kalika municipality, Chitwan, IDI***

#### Theme 2: Measles-rubella surveillance core functions

##### i. Case detection

According to the district and municipal respondents, doctors, health workers working in health facilities play a main role in detecting a measles case. Many respondents from the district, municipality, and both reporting and NRUs were not aware about standard case definition (SCD). In addition to the fever and rash, they also mentioned coryza, red, watery eyes, runny nose, and cough under SCD. A few of them mentioned koplik’s spots as the confirming sign of measles.

> *“Red rashes behind the mouth that went through the ear to the back, and sudden fever and cough, in child aged between 9 and 15 months is considered a suspected measles case”*
>
> ***-District level respondent, KII***
>
> *“Ear pain, followed by small red spots, fever, cold and cough, and a body rash is considered for detecting a suspected measles case”*
>
> ***-RU respondent, Raksirang rural municipality, Makwanpur, IDI***

The WHO-SMOs said that they provide support in detecting cases through phone or video calls when it becomes challenging for any health worker. All the respondents from the district and municipality reported that SCD were not posted at their institutions. Whereas, many respondents from the RU health facilities and few from NRUs mentioned that it was posted in their outpatient departments (OPDs) which was also verified through observations (Table 3).

> *“We have SCD posters for suspected measles-rubella at our health facility*.*”*
>
> ***-RU respondent, Bharatpur metropolitan city, Chitwan, IDI***

##### ii. Case notification

All the respondents from districts and RUs mentioned that notification occurs immediately upon detection of suspected measles cases. All the district respondents mentioned that the notification involves health facilities notifying the municipality and WHO-SMO, which then notifies the district. The municipal respondents also agreed that when the case is notified to the municipality, they further notify the district.

All respondents from RUs mentioned that they are responsible to notify the suspected measles cases. Many of them notified the cases to WHO-SMO while few reported that they also notified municipalities. WHO-SMOs also agreed that the RUs notify them when a suspected measles case is detected. They also added that if the RUs have not notified the district, then WHO-SMOs notify the district. Respondents from the district, municipal and RUs mentioned that the notification mainly occurs through letters, emails, SMS or group messages, Viber and WhatsApp. Few respondents from the non-reporting health facilities also said that they notify the suspected measles cases to the WHO-SMOs.

> *“The case notification involves health facilities notifying the municipality and WHO-SMO, which then notifies the district*.*”*
>
> ***-District level respondent, KII***
>
> *“Notifications are done through phone calls, SMS or group messages”*
>
> ***-Municipal level respondent, Chitwan, KII***

##### iii. Case investigation

Many of the district respondents mentioned that there is no guideline for case investigations. Only some of them reported that the district immunization focal person is responsible for the case investigation. A few mentioned that they mobilize teams and resources immediately to collect samples and visit health facilities for investigation.

At the municipal level, many respondents mentioned that the health section chief is responsible for leading the case investigation. A few also mentioned that in charge of health facilities were responsible for case investigation.

> *“There is no dedicated focal person assigned for case investigation. Mostly, we proceed under the leadership of the health section chief*.*”*
>
> ***-Municipal level respondent, Chitwan, KII***

Some of the respondents from the RU also reported that there is no dedicated focal person responsible for case investigation. Some of them reported that case-investigation forms (CIFs) are available and are filled at their health facilities. Some of the respondents from the NRUs mentioned that the district focal person is responsible for case investigation.

Neither municipal level respondents nor respondents from RUs were aware about the separate guidelines or standard operating procedure for case investigations. Both of these respondents mentioned that the case investigation is carried out based on the VPD surveillance orientation and onsite coaching conducted by WHO-SMOs.

WHO-SMOs mentioned that the case investigation is carried out in coordination among a team consisting of the RU health facilities in-charge, doctors, and district immunization focal person.

##### iv. Sample collection and transportation

Many respondents from the districts and municipalities were not aware about the timeframe for the sample collection from suspected measles cases. They mentioned that sample collection occurs at health facilities and is then sent to the WHO-IPD field office.

All the respondents from the RU mentioned that a blood sample is collected immediately for laboratory testing, when a patient with fever and rash arrives at a health facility. They mentioned that after collection, the sample is transported to the WHO-IPD field office while maintaining the reverse cold chain. WHO-SMO mentioned that, once the WHO-IPD field office receives the sample, it is sent to designated laboratories for testing. They reported that the WHO-IPD provides the transportation cost of shipment of samples and travel allowance to the health workers who are shipping the sample to the field office. The WHO-SMOs mentioned that when they receive the lab results from the designated laboratories, they share the results with health facilities within 4-5 days of sample shipment. A respondent from a RU mentioned that sample shipment can sometimes be delayed by 2-4 days due to the absence of responsible health workers for dispatching the sample.

> *“If there is a staff shortage in our health facility to ship the collected samples, it takes time……we manage to deliver it within two to four days*.*”*
>
> ***-RU respondent, Barhabise municipality, Sindhupalchok, IDI***

One of the respondents from the RU mentioned that they have neither the skills nor knowledge to collect samples and its shipment, thus, are referring the cases to other health facilities. Some rural health facilities mentioned the challenge of sample transportation due to difficult geographical terrain.

##### v. Reporting

All the district and municipal respondents were aware that weekly reporting of VPD cases is done by the RU health facilities. However, they are not aware of the status of timeliness and completeness of the weekly reporting from the health facilities, as they do not receive the weekly reports.

> *“RUs do not send weekly reports directly to the district health office, so we are not sure about the status of the timeliness and completeness of weekly reporting”*
>
> ***-District level respondent, KII***

Many respondents from the RUs said that they have been submitting weekly reports to WHO-SMO. However, they mentioned that sometimes delays in timely weekly reporting can occur if the person responsible is on leave or due to administrative issues, such as the unavailability or transfer of the chief officer who needs to sign the report. They mentioned that the weekly reports are sent through phone messenger or Viber to WHO-SMO and documented in the VPD registers. WHO-SMOs also mentioned that there is completeness in the weekly reporting but there have been some delays in timely reporting when there is absence of a focal person in the RU health facilities. This was also supported by the observation done at the RU health facilities. Many RUs reported no challenges in completeness of weekly reporting over the past six months. WHO-SMO mentioned that, when they are not able to visit the health facilities for a month, few health facilities would think that the VPD surveillance program has been closed and they don’t have to weekly report the cases.

> *“We can’t reach all the places every month. In some places if we don’t go for a month, they think that the VPD surveillance program has been closed and they don’t have to report*.*”*
>
> ***-WHO-SMO, IDI***

##### vi. Data analysis and utilization

All the respondents from RUs said that they rely on WHO-SMOs for data analysis and feedback on VPD surveillance. WHO-SMOs also mentioned they analyze the surveillance data on a monthly basis and share it to FWD, province, district through WHO-IPD main office. Some respondents from the RUs said that due to the small number of cases in their respective areas, they do not find data analysis as important for the measles surveillance in their community. Some of the respondents at all levels said that they use the analyzed data for planning health related activities.

##### vii. Review meetings

Municipality respondents reported that WHO-SMO conducts the review meetings for VPD surveillance in coordination with the district. Some municipalities and RU respondents reported that they have set up their own provision of additional monthly review meetings where activities related to surveillance are also included.

> *“WHO-SMO conducts the review meeting. The district plays a coordinating role and health workers from the RUs of different municipalities participate in the review meeting”*.
>
> ***-Municipal level respondent, Chitwan, KII***

#### Theme 3: Outbreak and response

The district and municipal respondents reported their role in the measles outbreak preparedness and response as planning, coordination with the health workers and WHO-SMO, and mobilization of the RRT. The municipal respondents mentioned that they were also responsible for supporting the health facilities in case investigation, sample collection and transportation, case management, conducting Outbreak Response Immunization (ORI) and providing Vitamin A supplementation during the outbreak preparedness and response.

> *“When the district is notified about the outbreak from the health facility and WHO-SMO, we communicate with RRT and mobilize them*.*”*
>
> ***-District level respondent, KII***

District respondents mentioned that they have a District Disaster Management Committee, chaired by the Chief District Officer to support the outbreak response. Both the district and municipality have their own functional Rapid Response Team (RRT) to support the outbreak response. Similarly, the respondents from both the RU and NRUs mentioned that they coordinate with the district and municipality for outbreak preparedness and response. Respondents from the RU described their role in identifying and reporting suspected measles cases to higher authorities, and in supporting sample collection and transportation. Many respondents from the RU mentioned that RRT played a major role during contact tracing. However, few of them reported that support from RRT was not received during the measles outbreak response.

Respondents from the NRUs mentioned that they were responsible for referring suspected measles cases to hospitals with laboratory facilities. WHO-SMOs reported their roles as providing support in case detection, case investigation, sample collection, and transportation during outbreak preparedness and response.

When asked about availability of operational guidelines for the outbreak preparedness and response, none of the respondents from any levels were aware of any such guidelines. However, WHO-SMOs mentioned that there is an existing VPD surveillance guideline which includes the standard operating procedure for outbreak preparedness and response.

> *“We don’t have guidelines related to outbreak preparedness and response”*
>
> ***-RU respondent, Helambu rural municipality, Sindhupalchok, IDI***

Many respondents in both RUs and NRUs were not aware about the definition of the suspected measles outbreak. When asked about a confirmed measles outbreak in the last three years to respondents from all levels, only one municipality had recorded a measles outbreak last year. The respondents from the municipality which experienced the outbreak shared that the outbreak response was effective and it was possible by good coordination and support from WHO, higher public health authorities and the RRT team.

> *“At that time, initially, two children with fever and rash came to the health facility. In the health facility, the health workers (doctors) examined the children and identified them as suspected cases. Samples were taken from those two children and sent to laboratory test for ELISA. It was confirmed as positive. Following that, health workers were deployed to the children’s home area. They found 8 to 10 more children with similar symptoms, and there was also one pregnant woman with similar symptoms. Among them, samples were collected from five children, were taken, and sent for lab tests. Their results also came as positive. It was considered an outbreak, and it was managed accordingly. Investigation continued, and contact tracing was conducted in the village. They looked for contacts, provided Vitamin A, and conducted awareness activities in the field. Later, in coordination with WHO and our public health office, it was decided to conduct an Outbreak Response Immunization (ORI). The ORI was carried out as part of the outbreak management*.*”*
>
> ***-Municipal level respondent, Chitwan, KII***

Regarding the communication channels during outbreak preparedness and response, all respondents mentioned that communication is maintained among all levels through phone calls or text from social media like facebook groups or letters or mails.

When asked about challenges in outbreak preparedness and response, respondents from private health facilities working as RUs mentioned that they have limited resources and the government of Nepal provides no support with supplies to manage outbreaks. On top of that, the government of Nepal expects to implement their program free of cost.

#### Theme 4: Measles-rubella surveillance supportive functions

##### i. Roles and responsibilities in measles-rubella surveillance

District and municipal respondents reported that their responsibilities primarily include coordination, monitoring, and supervision of overall health services, including VPD surveillance. Respondents from RUs mentioned that they are responsible for vaccination and surveillance programs. WHO-SMOs mentioned that their key role is to orient health workers in both public and private facilities on VPD surveillance. They added that their roles included weekly visits to high-priority reporting sites, conducting active surveillance to ensure no cases are missed, and coordinating sample transportation.

- **Coordination for measles-rubella surveillance** All the district respondents reported that coordination for measles–rubella surveillance is done at all levels (province, municipalities and WHO-IPD). They reported that they coordinate with the municipality in terms of supporting the vaccination programs, measles-rubella outbreak investigation and response. Similarly, with the WHO-IPD, they coordinate to ensure necessary logistics (recording and reporting tools, checklists and specimen collection equipment) for surveillance activities at the RUs. They further mentioned that WHO-SMO acts as a liaison between the district and the local level for VPD surveillance activities.

The municipal respondents reported that the municipality coordinates with the district health office for VPD surveillance activities. Many respondents from reporting and NRUs said that they coordinate with the district and municipality for resources and technical support. Further, respondents from municipal level, reporting and NRUs said that they coordinate with WHO-SMO in case of an outbreak.

> *“Coordination with WHO-SMO is very good. We reach out to them when needed, often through messenger, and they are easy to contact*.*”*
>
> ***-RU respondent, Bharatpur metropolitan city, Chitwan, IDI***

WHO-SMOs reported that they coordinate with all levels of government in case of measles outbreak response. They mentioned that they have been coordinating with the government bodies (district and municipalities) to engage them to ensure ownership of the surveillance program. They also mentioned that they coordinate with the district, municipalities and their RRT to support case investigation of suspected cases, including active search during the measles outbreak and ORI.

> *“Upon receiving a case notification from a RU, I coordinate with the district and local government. For suspected measles cases and outbreak, I coordinate with the municipality and district RRT team, including WHO-IPD and the Family Welfare Division. I support the RRT for active search and for sample collection and transportation. In the event of an outbreak, coordination with the Family Welfare Division is essential for ORI and vaccine distribution*.*”*
>
> ***-WHO-SMO, IDI***

They also mentioned that with the FWD and WHO-IPD, they coordinate to notify about the suspected measles cases. They also coordinate with the NPHL for sample transportation and receiving laboratory reports. Upon receiving the lab reports, they further coordinate with FWD and WHO-IPD and respective health facilities in communicating the laboratory results (positive or negative).

- **Monitoring of the measles-rubella surveillance** All the districts and municipality respondents reported that they were responsible for the monitoring of the VPD surveillance including measles-rubella. District respondents reported that they monitor the availability of necessary surveillance tools and sample collection equipment provided by WHO at the reporting sites. However, some of the district respondents reported that they are also responsible for several health programs in addition to the VPD surveillance so the regular monitoring is quite challenging. They conduct monitoring jointly with WHO-IPD in case of measles outbreak. Few respondents from districts described that although they do not have specific monitoring activity dedicated towards VPD surveillance, they conduct integrated monitoring visits for overall health programs.

> *“We are conducting integrated monitoring. Not only surveillance but overall health programs are being monitored*.*”* - ***District level respondent, KII***

- **Supportive supervision** Respondents from district and municipality reported that specific supervision for VPD surveillance activities was not conducted, with most supervision being integrated with immunization and nutrition programs. The frequency of supervision varied: district respondents reported monthly visits, while municipal respondents visits every two to three months, and in some cases, only once a year. Due to the challenges in reaching all health facilities monthly for supervision, both district and municipal respondents reported that they conduct supportive supervision in a turn wise manner i.e. they visit one health facility at one time and for next visit they plan another health facility.

> *“According to our plan, there is a provision that at least one health facility should be visited for supervision at least three or four times a year, i*.*e. quarterly. However, since there are many health facilities in our district, it is quite difficult to reach all the facilities*.*”*
>
> ***-District level respondent, KII***

One of the municipal respondents mentioned that feedback on surveillance and vaccination performance status were provided during supervision visits. Many respondents reported that there are no significant challenges regarding supervision of surveillance, but some said that the staffing is not adequate to conduct regular supervisions in a timely manner.

##### ii. Training on VPD surveillance

Many respondents from districts and municipalities mentioned they have not received specific training on VPD surveillance. WHO-SMOs explained that they organize orientation programs on VPD surveillance for health workers but not formal training. These orientations are conducted at both district and sub-district levels. The orientation covers topics like case detection, notification, case investigation forms, sample collection, and reporting completeness and timeliness. However, some respondents from RUs mentioned they have not received any orientation or training related to VPD surveillance. Some of them mentioned that there is training being conducted on vaccination campaigns which also includes sessions on VPD surveillance.

> *“I haven’t received any specific training for VPD surveillance. During vaccination campaigns, we get training including sessions on VPD surveillance, but there isn’t any specific training for VPD surveillance*.*”*
>
> ***-RU respondent, Hetauda sub-metropolitan city, Makwanpur, IDI***

##### iii. Resources

- **Financial resources** District and municipal respondents mentioned that there is no specific budget allocation for VPD surveillance and outbreak response. All districts and municipality respondents reported they manage budgets for measles outbreaks and surveillance from budgets allocated and received under other areas such as disaster funds. Municipalities receive conditional grants for disaster management from the federal government.

> *“No specific budget is allocated for VPD surveillance*.*”*
>
> ***-District level respondent, KII***

- **Logistics and supplies**

Many respondents from all levels mentioned that WHO-IPD provides the necessary surveillance tools and sample collection supplies. Respondents from the RU health facilities mentioned that they have sufficient supplies for sample collection and transportation. WHO-SMOs mentioned that WHO-IPD promptly restocks items like gloves, blood collection vials, cryo vials, and butterfly cannulas when shortages are reported.

District respondents reported that they supply health facilities with necessary supplies such as ice packs, vaccine carriers, and essential medicines like Vitamin A and paracetamol. Overall, respondents indicated no significant logistical challenges for measles-rubella surveillance.

> *“WHO-IPD provides sufficient sample collection equipment and surveillance tools to all the reporting sites. Whenever we ask for the supplies, they provide us as per our request”*
>
> ***-RU respondent, Bharatpur metropolitan city, Chitwan, IDI***

- **Human resources** Many respondents from the district, municipalities, and RUs mentioned that the existing human resource for surveillance at their respective levels is sufficient. However, some respondents at the district and municipality levels indicated that there is no sanctioned position of focal person for VPD surveillance at all levels (health facility, municipality and district). Despite this, they reported that surveillance has been carried out using the existing human resource.

> *“We don’t have a dedicated human resource for VPD surveillance. We lack resources. We manage with the existing health workers*.*”*
>
> ***-Municipal level respondent, Sindhupalchok, KII***

WHO-SMO reported that the number of health workers in the RUs is generally sufficient, particularly in health facilities with low patient flow. Many respondents from RUs mentioned having an alternative focal person for surveillance in the absence of the designated one. However, WHO-SMO emphasized the need for more than one focal person to handle VPD surveillance activities in tertiary hospitals due to the high workload.

#### Theme 5: Experience and willingness of Rapid Diagnostic Test utilization

- **Past experience of using any RDT** All the district respondents said they have no experience of using any kind of RDT kits. Respondents from the municipality and RUs mentioned that they have had past experience of using RDT for COVID, kala-azar, malaria, dengue and HIV. Respondents who have used RDT kits in the past shared that they did not face any challenges while using it.

- **Willingness to use measles RDT in future** Almost all of the respondents were willing to use RDT Measles in the future if it were to be made available. Few respondents from the NRUs preferred measles RDT which does not require withdrawing blood samples. When asked about the advantage of measles RDT use, some of the respondents from RUs reported that it would also be easier to use measles RDTs in children due to challenges with withdrawing blood samples.

Overall, respondents mentioned that orientation on how to use the measles RDT would be helpful. They reported that it would be advantageous for timely detection, case investigation and case management. Furthermore, they mentioned that the public also wants timely results of their tests, so it might be helpful in increasing people’s satisfaction. Some of them said that they cannot fully depend only on measles RDT results and the existing laboratory confirmation of the measles cases should be run concurrently. One of the WHO-SMOs said that they would recommend using measles RDT only after it is tested for its validity and reliability as they are more prone to false positives and false negatives results.

> *“If there’s an outbreak, use of RDT can be helpful for timely detection of the disease, case management and prevent further spread*.*”*
>
> ***-RU respondent, Raksirang rural municipality, Makwanpur, IDI***

## Discussion

Our findings revealed that the key strategy to measles elimination which is achieving MR2 coverage of 95% was not achieved in most of the municipalities. Studies suggest that none of the WHO regions had achieved the target of measles elimination by the end of 2019.^22-23^ In our study, the completeness and timeliness of health facilities weekly reports in the previous six months was 95% and 55% respectively. The timeliness is quite comparable with a study done in Indonesia where it was 50% and however completeness was only 59% (much lower than in our study).^24^ Further, a study done in India mentioned that timely and reliable information has been provided there through VPD surveillance system which has helped in delivering immediate public health response and forming evidence-guided policies and plans.^25^ The issue in completeness of patient data can create obstacles in contact tracing as well as locating cases.^26^ Likewise, the completeness of the weekly reported data if accurate, can provide valuable information to measure the disease burden in the community.^27^ The public health response to be delivered in the community depends upon the surveillance systems which provide timely, complete and reliable information as support for epidemiological investigation activities, hence this part of surveillance is truly crucial for moving towards disease elimination.^26^

Measles was mentioned as one of the VPDs under current VPD surveillance by most of the respondents from all levels in our study. The measles-rubella surveillance system under the Indonesia Health Ministry (IHM) also considers measles and rubella as potential outbreak diseases. Case based measles surveillance (CBMS) of Indonesia is used to detect the cases and respond to disease outbreaks earlier.^24^ Many participants from all levels in our study said that the case investigation is initiated immediately after case notification while only some of the respondents from RUs said that CIFs are available and filled at their health facilities. The study done in India reported that more than 90% of suspected cases are investigated within 48 hours of notification in India and the epidemiological data are collected from CIF.^25^ Many respondents from the RUs said that they have been submitting weekly reports to WHO-SMO through phone messenger or Viber to WHO-SMO and documented in the VPD registers. Study done in Indonesia stated that they maintained case-based surveillance reports on a daily basis whereas they too updated aggregate reports on a weekly basis. These weekly data in Indonesia is examined by DHO officer. They mentioned that they use social media to send weekly reports to their central server. Indonesia also has guidelines which for sending reports however, the study mentioned that each PHC there reports cases differently.^24^

Study done in Indonesia suggested human resource shortages in their PHCs which affected surveillance system to operate smoothly.^24^ Our findings also suggest no designated focal persons at health facilities, municipalities and district levels as one of the major barriers of VPD surveillance, implying that there is shortage of health workers in surveillance sector. Moreover, in our study, many district and municipal level respondents said that they have not received specific trainings on VPD surveillance except from orientation programs covering some topics of VPD surveillance like case detection, notification, case investigation forms, sample collection, and reporting completeness and timeliness. In contrast, a study done in India mentioned that they regularly conduct workshops at district as well as sub-district levels by using surveillance posters and other appropriate audio-visual aids.^25^ This is important for sensitizing the healthcare workers on the necessity of VPD surveillance and its overall process.^25^

District and municipal respondents from our study mentioned no specific budget allocation for VPD surveillance and outbreak response, although they do receive budget under disaster funds in general. This is supported by results of Indonesian study which also mentioned that they lacked standard budget allocation for measles-rubella disease control. This resulted in varying availability of budget under measles-rubella disease control in each PHC of Indonesia.^24^ Additionally, funding for the specimen transportation in Indonesia is limited which affects the PHC officers’ performances in sample transportation.^28^ Our findings suggested that transportation cost of sample shipment and travel allowance of health workers involved in shipment is provided by WHO-IPD in Nepal.

All the respondents from RUs in our study said that they rely on WHO-SMOs for data analysis and feedback on VPD surveillance, suggesting that they do not analyze data themselves. The data analysis in health facilities itself was found to be low in Indonesia as well.^24^ Study done in sub-Sahara Africa suggests that the quality of data analysis affects its usefulness^29^ hence data analysis is significant to be conducted in every level of facilities involved in surveillance.^27^ Nearly all respondents form all level in our study mentioned that VPD surveillance plays a crucial role in measles elimination. A study done in India stated that MR surveillance indeed is guiding their progress towards achievement of MR elimination in the country.^30^

## Conclusions

Although, national immunization programme (NIP) is a priority 1 (P1) programme of the Government of Nepal, one of the key strategies (95% MR2 coverage) to achieve measles elimination was not achieved in most of the municipalities. This study suggests that measles-rubella surveillance in the study sites can be strengthened through capacity building of health workers in VPD surveillance, ensuring the availability of the VPD surveillance field guide, availability of the dedicated focal person, emphasizing in data analysis and utilization of surveillance data.

## Ethics statements

### Patient consent for publication

Not applicable

### Ethical approval

Ethical approval was granted from the Nepal Health Research Council (NHRC) for the study (reference 240-2024). For the in-person interviews, a written informed consent was obtained from individuals who agreed to participate in the study and to record the interview.

The research team oriented on the ethical aspects of the study, including the emphasis on participants’ privacy, confidentiality, and anonymity both during and after the research. The voluntary nature of participation and the right to withdraw from the study at any time were also clearly communicated. An advance appointment was arranged from the participants with careful consideration to ensure that it does not interfere with their working hours and patient care time.

## Acknowledgement

We would like to Family welfare division (Department of Health Services), Gavi, the vaccine alliance, WHO-IPD, and technical working group members for providing necessary support in this work

## Contributors

SRU, BST, RS, DS, LK: Study design and conceptualization, data interpretation. BS, KK, SK, AU: Data acquisition, verification, analysis, interpretation. All authors were involved in drafting and critical revision of the manuscript. All authors approved the final manuscript. BS and SK are joint first authors.

## Data availability

Data will be made available upon reasonable request.

## References

1. World Health Organization. Measles [Internet]. [cited 2024 Sep 25]. Available from: https://www.who.int/news-room/fact-sheets/detail/measles

2. Minta AA, Ferrari M, Antoni S, Portnoy A, Sbarra A, Lambert B, Hatcher C, Hsu CH, Ho LL, Steulet C, Gacic-Dobo M, Rota PA, Mulders MN, Bose AS, Caro WP, O’Connor P, Crowcroft NS. Progress Toward Measles Elimination — Worldwide, 2000–2022. Morbidity and Mortality Weekly. 2023;72(46):1262–1268. Available from: https://www.cdc.gov/mmwr/volumes/72/wr/mm7246a3.htm

3. Family Welfare Division, Government of Nepal. Technical Report on Measles Outbreaks and Root Cause Analysis, Nepal, 2022–2023 [Internet]. Available from: https://fwd.gov.np/wp-content/uploads/2024/02/Technical-reports-of-Root-Cause-Analysis-and-Measles-Outbreak-2022-23.pdf

4. World Health Organization. Global Measles and Rubella Strategic Plan 2012-2020 [Internet]. 2012. Available from: https://iris.who.int/bitstream/handle/10665/44855/9789241503396_eng.pdf

5. CDC. Global Measles Vaccination. 2024 [cited 2024 Sep 25]. About Global Measles. Available from: https://www.cdc.gov/global-measles-vaccination/about/index.html

6. World Health Organization. Immunization coverage [Internet]. [cited 2024 Sep 25]. Available from: https://www.who.int/news-room/fact-sheets/detail/immunization-coverage

7. Sadaf A, Richards JL, Glanz J, Salmon DA, Omer SB. A systematic review of interventions for reducing parental vaccine refusal and vaccine hesitancy. Vaccine. 2013 Sep 13;31(40):4293–304.

8. World Health Organization. Strategic plan for measles elimination and rubella and congenital rubella syndrome control in the South-East Asia Region, 2014-2020 [Internet]. [cited 2024 Sep 25]. Available from: https://iris.who.int/handle/10665/205923

9. World Health Organization, Regional Office for South-East Asia. Measles and Rubella Elimination by 2023 [Internet]. Available from: https://apps.who.int/iris/bitstream/handle/10665/327923/sea-rc72-r3-eng.pdf?sequence=1&isAllowed=y

10. World Health Organization. Review of progress and way forward on measles and rubella elimination activities in the WHO South-East Asia Region.

11. Crowcroft NS, Minta AA, Bolotin S, Cernuschi T, Ariyarajah A, Antoni S, et al. The Problem with Delaying Measles Elimination. Vaccines. 2024 Jul 22;12(7):813.

12. WHO Regional Office for South-East Asia. South-East Asia Region EPI Factsheet 2024. 2024.

13. Asia WHORO for SE. Regional Consultation on resetting the target date for achieving the goal of measles and rubella elimination in the WHO South-East Asia Region, New Delhi, India, 14–16 March 2023. 2023 [cited 2024 Sep 25]; Available from: https://iris.who.int/handle/10665/368018

14. World Health Organization. Strategic plan for measles and rubella elimination in WHO South-East Asia Region: 2020–2024 [Internet]. [cited 2024 Sep 25]. Available from: https://iris.who.int/handle/10665/330356

15. World Health Organization. More than 1.5 Million Children Vaccinated Against Measles and Rubella in Nepal [Internet]. [cited 2024 Sep 25]. Available from: https://www.who.int/nepal/news/detail/19-07-2023-more-than-1.5-million-children-vaccinated-against-measles-and-rubella-in-nepal

16. Department of Health Services, Government of Nepal. Annual Report 2022/2023 [Internet]. Available from: https://dohs.gov.np/wp-content/uploads/Annual_Report.pdf

17. IA2030. Immunization Agenda 2030 [Internet]. [cited 2024 Sep 25]. Available from: https://www.immunizationagenda2030.org/

18. Kumar S, Lennon P, Uranw S, Fielding T, Mvundura M, Drolet A, et al. Using freeze-preventive cold boxes in rural Nepal: A study of equipment performance, acceptability, system fit, and cost. Vaccine X. 2024 Feb 28;18:100467.

19. World Health Organization. Immunization Preventable Diseases Database. 2022.

20. Khanal S. Progress Toward Measles Elimination — Nepal, 2007–2014. Morbidity and Mortality Weekly Report [Internet]. 2016 [cited 2024 Sep 25];65. Available from: https://www.cdc.gov/mmwr/volumes/65/wr/mm6508a3.htm

21. Rachlin A, Hampton LM, Rota PA, Mulders MN, Papania M, Goodson JL, et al. Use of Measles and Rubella Rapid Diagnostic Tests to Improve Case Detection and Targeting of Vaccinations. Vaccines. 2024 Aug;12(8):823.

22. N.C. Makova, et al. Evaluation of the measles case-based surveillance system in Kwekwe city, 2017-2020: a descriptive cross-sectional study Pan African Med J, 42 (May 2022), 10.11604/pamj.2022.42.113.31373

23. Khanal S, Kassem M.A., Bahl S. Progress toward measles elimination inSouth-east Asia region, 2003–2020. Morbidity and Mortality weekly report [Internet]. https://www.ncbi.nlm.nih.gov/pmc/articles/PMC9400531/

24. Sylvia Gusrina, Mei Neni Sitaresmi, Samsu Aryanto, Bayu Satria Wiratama, Evaluation of measles-rubella control and prevention program implementation: System and community review. Clinical Epidemiology and Global Health. 2024;29 101758, ISSN 2213-3984. 10.1016/j.cegh.2024.101758

25. Kumar A, Murugan R, Donkatti S, Sharma D, Kaundal N, Avagyan T, Kumar P, Bahl S, Khanal S, Bura V. Strengthening of vaccine-preventable disease (VPD) surveillance to enhance national health capacity and security: perspective from India. 2024;12(8):941. https://www.mdpi.com/2076-393X/12/8/941

26. Kalil FS, Bedaso MH, Abdulle MS, Mohammed NU. Evaluation of measles surveillance systems in Ginnir district, Bale zone, southeast Ethiopia: a concurrent embedded mixed quantitative/qualitative study. Risk Manag Healthc Pol. 2021;14:997–1008. 10.2147/RMHP.S295889

27. Alemu T, Gutema H, Legesse S, Nigussie T, Yenew Y, Gashe K. Evaluation of public health surveillance system performance in Dangila district, Northwest Ethiopia: a concurrent embedded mixed quantitative/qualitative facility-based cross-sectional study. BMC Public Health. 2019 Oct;19(1). 10.1186/s12889-019-7724-y

28. Kusumawardani EF, Laily SR, Sipahutar RY, Domingga M, Martini S. Evaluation of Measles Surveillance System in Provincial Health Office, East Java, Indonesia. 2020. https://medic.upm.edu.my/upload/dokumen/2020010214592508_MJMHS_0088.pdf

29. Mremi IR, George J, Rumisha SF, Sindato C, Kimera SI, Mboera LE. Twenty years of integrated disease surveillance and response in Sub-Saharan Africa: challenges and opportunities for effective management of infectious disease epidemics. One Health Outlook. 2021 Dec;3(1). 10.1186/s42522-021-00052-9

30. Ministry of Health and Family Welfare, Government of India. Introduction of Measles Rubella Vaccine (Campaign and Routine Immunization). National Operational Guidelines. Government of India. New Delhi, India, 2017. https://www.mohfw.gov.in/sites/default/files/195431585071489665073.pdf

